# Intervention Serology and Interaction Substitution: Modeling the Role of ‘Shield Immunity’ in Reducing COVID-19 Epidemic Spread

**DOI:** 10.1101/2020.04.01.20049767

**Authors:** Joshua S. Weitz, Stephen J. Beckett, Ashley R. Coenen, David Demory, Marian Dominguez-Mirazo, Jonathan Dushoff, Chung-Yin Leung, Guanlin Li, Andreea Măgălie, Sang Woo Park, Rogelio Rodriguez-Gonzalez, Shashwat Shivam, Conan Zhao

## Abstract

The COVID-19 pandemic has precipitated a global crisis, with more than 690,000 confirmed cases and more than 33,000 confirmed deaths globally as of March 30, 2020 [1–4]. At present two central public health control strategies have emerged: mitigation and suppression (e.g, [5]). Both strategies focus on reducing new infections by reducing interactions (and both raise questions of sustainability and long-term tactics). Complementary to those approaches, here we develop and analyze an epidemiological intervention model that leverages serological tests [6, 7] to identify and deploy recovered individuals as focal points for sustaining safer interactions via interaction substitution, i.e., to develop what we term ‘shield immunity’ at the population scale. Recovered individuals, in the present context, represent those who have developed protective, antibodies to SARS-CoV-2 and are no longer shedding virus [8]. The objective of a shield immunity strategy is to help sustain the interactions necessary for the functioning of essential goods and services (including but not limited to tending to the elderly [9], hospital care, schools, and food supply) while decreasing the probability of transmission during such essential interactions. We show that a shield immunity approach may significantly reduce the length and reduce the overall burden of an outbreak, and can work synergistically with social distancing. The present model highlights the value of serological testing as part of intervention strategies, in addition to its well recognized roles in estimating prevalence [10, 11] and in the potential development of plasma-based therapies [12–15].

---

In the absence of reliable pharmaceutical interventions against SARS-CoV-2, multiple public health strategies are being deployed to slow the coronavirus pandemic [1, 5, 16]. These strategies can be broadly grouped into two approaches: mitigation; and suppression. Mitigation includes a combination of social distancing (including school and university closures), case testing, and symptomatic case isolation to reduce epidemic spread and burden on hospitals. Mitigation is intended to lessen an outbreak, however the level of disease may still overwhelm health services [5]. Instead, some jurisdictions have either pre-emptively or reactively adopted a combination of travel restrictions (shown to be effective in curtailing dispersion if implemented early enough [17, 18]) and suppression: imposing complete shut-downs of the bulk of non-essential services for extended periods (e.g., sheltering in place). Suppression has led to marked decreases in prevalence in the short term by combining case isolation, quarantine, use of separate facilities for treating COVID-19 patients, and large-scale viral testing to reduce transmission. Suppression also comes with significant costs, threatening social order and socio-economic health.

Here, we propose a complementary intervention approach that is intended to reduce transmission while lessening the costs of suppression and mitigation. The core idea is to leverage a mechanism of ‘interaction substitution’ by identifying and deploying recovered individuals who have protective antibodies to SARS-CoV-2. The intent is to develop population-level ‘shield immunity’ by amplifying the proportion of interactions with recovered individuals relative to those of individuals of unknown status (see Figure 1). Here, we assume that recovered individuals (i.e., virus-negative and antibody-positive) can safely interact with both susceptible and infectious individuals, in effect substituting interactions between susceptible and infectious individuals for interactions with a recovered individual. The intervention strategy is both local in scope and scales with outbreak size, given that the potential impact of shield immunity grows with a local outbreak. We recognize that our assumptions about safety for both recovered individuals and those they interact with is of vital importance. We return to this issue in discussing translational efforts of shield immunity.

**FIG. 1:**
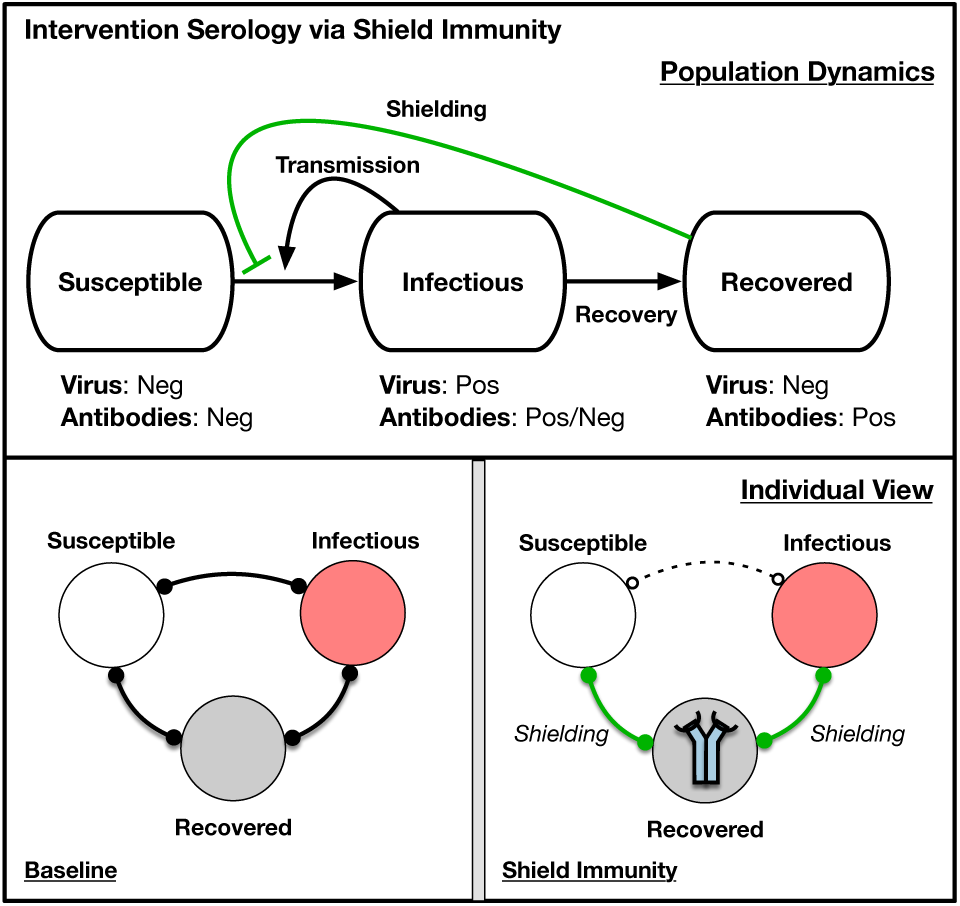
Simplified schematic of intervention serology via shield immunity. (Top) Population dynamics of susceptible, infectious, and recovered in which recovered individuals reduce contact between susceptible and infectious individuals. Arrows denote flows between population level-compartments. (Bottom) Individual view of baseline scenario and shielding scenario, in which the identification, designation, and deployment of recovered individuals is critical to enabling S-R and I-R interactions to replace S-I interactions. Bonds denote interactions between individuals. In the <u>Shield Immunity</u> panel, the icon in the recovered individuals denotes the identification of individuals with protective antibodies, and hence the enhanced contribution of such individuals to shield immunity in contrast to the <u>Baseline</u> panel. shield immunity.

To illustrate the concept of shield immunity, consider an epidemic model in which individuals tend to substitute their interactions with identified (or strategically located) recovered individuals. Hence, rather than mixing at random, we consider a relative preference of 1 + *α* that a given individual will interact with a recovered individual in what would otherwise be a potentially infectious interaction. The dynamics of the fraction of susceptible *S*, infectious *I*, and recovered *R* individuals are:

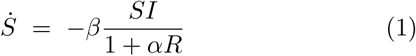

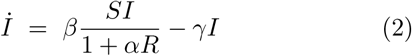

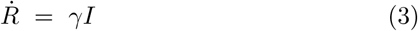

such that when *α* = 0 we recover the conventional SIR model. Note that the denominator of 1 + *αR* can be thought of as *S* + *I* + *R* + *αR*. Given that *S* + *I* + *R* = 1, this is equivalent to the term 1 + *αR*. Detailed mixing and substitution models could lead to variations of this model, e.g., in spatially explicit domains, on networks, etc [19–22]. Figure 2 illustrates shield immunity impacts on an SIR epidemic with ℛ_*0*_ = 2.5. In this SIR model, shield immunity reduces the epidemic peak and reduces epidemic duration. In effect, shielding acts as a negative feedback loop, i.e., given that the effective reproduction number is ℛ_*eff*_ (*t*)= ℛ0 = *S*(*t*)/(1 + *αR*(*t*)). As a result, interaction substitution increases as recovered individuals increase in number and are identified. For example, in the case of *α* = 20, the epidemic concludes with less than 20% infected in contrast to the final size of approximately 90% in the baseline scenario without shielding.

**FIG. 2:**
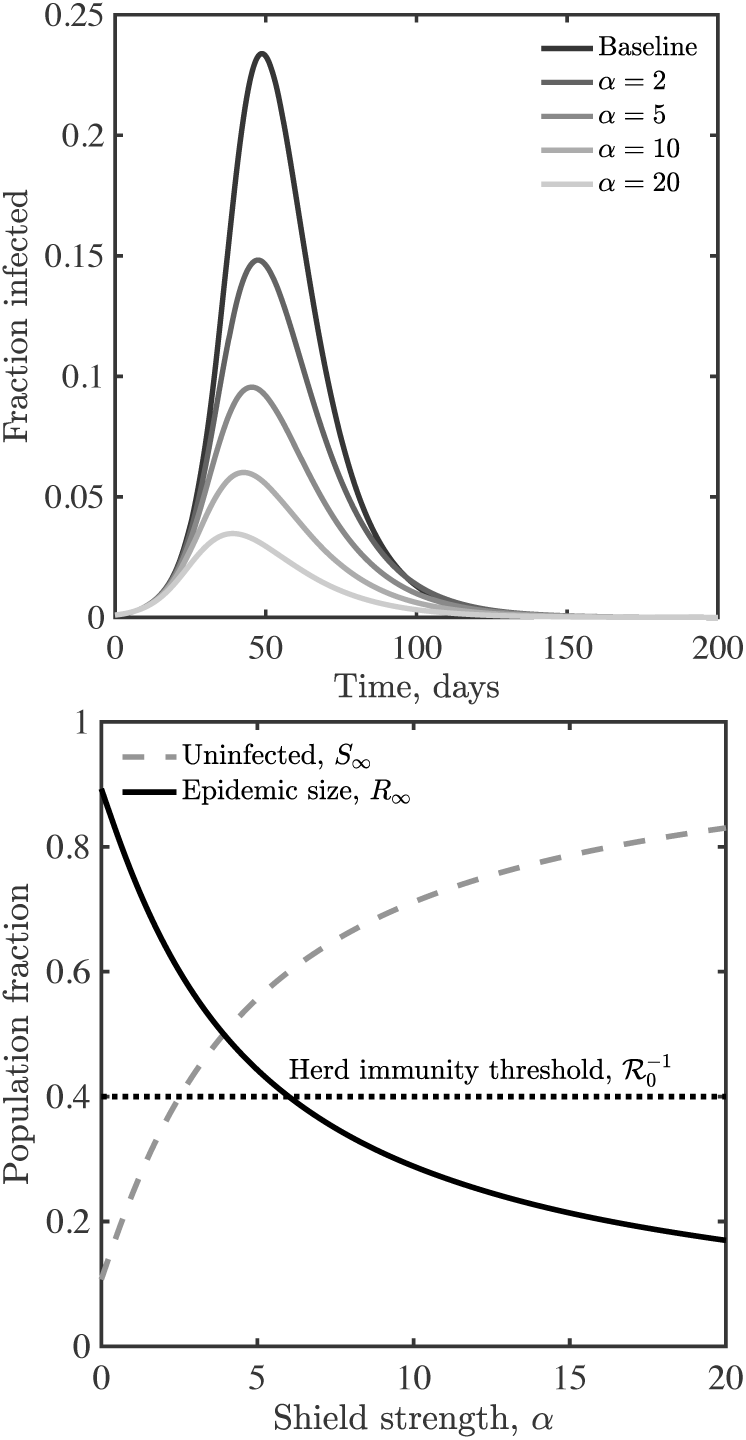
Shield immunity dynamics in a SIR model. (Top) Infectious case dynamics with different levels of shielding, *α*. (Bottom) Final state of the system as a function of *α*. In both panels, *β* = 0.25 and *γ* = 0.1.

We next apply the concept of shield immunity to the epidemiological dynamics of the COVID-19 pandemic, ignoring births and other causes of deaths for simplicity. Consider a population of susceptible *S*, exposed *E*, infectious asymptomatically *I*_*a*_, infectious symptomatically *I*_*s*_, and recovered *R* who are free to move, without restrictions in a ‘business as usual’ scenario. A subset of symptomatic cases will require hospital care, which we further divide into subacute *I*_*hsub*_, and critical/acute (i.e., requiring ICU intervention) *I*_*hcri*_ cases. We assume that a substantial fraction of critical cases will die. Age-stratified risk of hospitalization and acute cases are adapted from the Imperial College of London report [5]. The full model incorporating shield immunity (see SI for equations and details, and Figure S1 for a schematic) differs from conventional SIR models with social distancing or case isolation interventions in a key way: the rate of transmission is reduced by a factor of 1/(*N*_*t*_ + *αR_shields_* where *N*_*tot*_ denotes the fraction of the population in the ‘circulating baseline’, and *R*_shields_ denotes the total number of recovered individuals between the ages of 20-60 (a subset of the total recovered population). In this model, we assume that all recovered individuals have immunity, but that only a subset are available to facilitate interaction substitutions. The model assumes that the circulating pool is not interacting with hospitalized patients, which must be incorporated into implementation scenarios with healthcare workers (HCW-s), who represent an intended target for shield immunity [23]). The baseline epidemiological parameters, age stratified risk, and population structure are listed in the SI (adapted from [5, 24–27]; see github for code and full implementation details).

We use the baseline epidemiological parameters and seed an outbreak with a single exposed individual until the outbreak reaches 0.1% total prevalence, e.g., 10,000 individuals infected out of a population of 10,000,000, at which point a shielding strategy is implemented. Outbreak scenarios differ in transmission rates, with *ℛ*_0_ =1.57 and 2.33 in the low and high scenarios, respectively. Early estimates of *ℛ*_0_ from Wuhan are consistent with a 95% CI of between 2.1 and 4.5 [28], putting our high scenario on the conservative end of estimated ranges. However, the *ℛ*_0_ of the high scenario we examine here is consistent with the range of 2.0 to 2.6 considered by the Imperial College London group [5], and with the median of *ℛ*_*eff*_ = 2.38 (95% CI: 2.04-2.77) as estimated via stochastic model fits to outbreak data in China that accounts for undocumented transmission [26]. Moreover, control measures reduce transmission, and our low scenario is consistent with estimates of *ℛ*_*eff*_ = 1.36 (95% CI: 1.14-1.63) in China from Jan. 24 to Feb. 3 after travel restrictions and other control measures were imposed. Figure 3 shows the results of comparing interventions to the baseline case. As in the simple SIR model, shielding (on its own) could potentially decrease epidemic burden across multiple metrics, decreasing both the total impact and shortening the peak event. In a population of size 10,000,000 for the high scenario, the final epidemic predictions are 71,000 deaths in the baseline case vs. 58,000 deaths given intermediate shielding (*α* = 2), and 20,000 deaths given enhanced shielding (*α* = 20). In a population of size 10,000,000 for the low scenario, the final epidemic predictions are 50,000 deaths in the baseline case vs. 34,000 deaths given intermediate shielding, and 8,300 deaths given enhanced shielding. The majority of deaths are in those ages 60 and above, despite the lower fraction of individuals in those ranges (see Figure 3), consistent with estimates in related COVID-19 models [5, 25, 26] and from outbreaks in Italy and China [29]. Note that our simulation results consider impacts based on shielding alone; whereas ongoing restrictions via social distancing and shelter in place orders will reduce interaction rates (a point we revisit later). The effectiveness of shielding depends on the product of the number of potential shields identified and their effective substitutability, i.e, *αR*, combining identification of and interaction rate by shields.

**FIG. 3:**
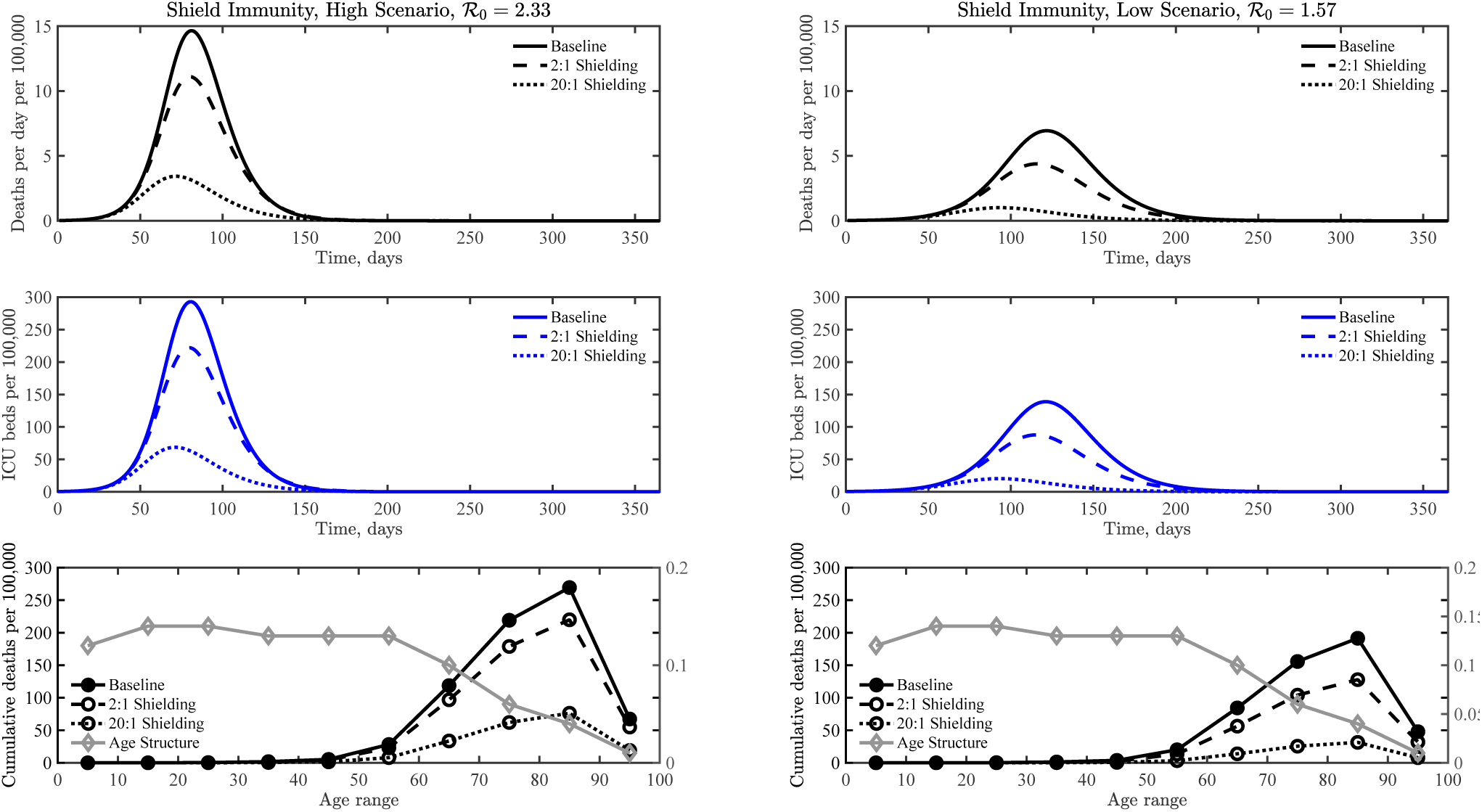
COVID-19 dynamics in a baseline case without interventions compared to two shield immunity scenarios, *α* = 2 and *α* = 20, including deaths, ICU beds needed, and age distribution of fatalities. See the SI for more details on alternative scenarios, high (left) and low (right).

The population-scale impacts of shielding depends on multiple factors, including demographic distributions, the fraction of asymptomatic transmission ([24, 26]), and the duration of immunity. The SI treats each of these items at length. First, we find that populations with a strongly right-shifted demographic distribution will receive more potential benefits from shielding. Even though there are fewer recovered individuals between the ages of 20-60 to draw from (in a relative sense), the impact of shield immunity is greater. We find that the relative reduction in deaths via shield immunity is proportional to the relative differences in the fraction of population over 60 (e.g., see SI for details on US-state level analysis, similar results hold for countries like Italy where more than 23% of the population is older than 65 and nearly 30% is older than 60; Figures S2-S3). Second, shield immunity is robust to variation in asymptomatic infection probabilities, improving outcomes in models with varying baseline levels of asymptomatic transmission (see Figures S4-S6). In addition, the present model results assumes that immunity has relatively fast onset and is permanent in duration. Clinical work in Zhejiang University, China, suggests that seroconversion of total antibody (Ab), IgM and IgG antibodies developed with a median period of 15, 18, and 20 days, respectively (albeit for symptomatic patients in a hospital; similar data for seroconversion of asymptomatic individuals was not included [30]). In the SI we show that impacts of shield immunity is robust insofar as the duration of immunity is 4 months or longer (Figures S7-S8); we note that distinct control measures that extend the epidemic would likely impact effectiveness of shield immunity. For context, the titer of protective antibodies in individuals infected with related betacoronaviruses (causing mild/moderate symptoms) reduced over a one year period such that re-exposure can lead to re-infection [31], in contrast to evidence of multi-year immunity for individuals recovered from SARS [32]. In addition, we emphasize that the accuracy of serological tests is key. The benefits of shield immunity can be undermined if recovered individuals can be re infected (even with little danger to them), or potentially misidentified, leading to interaction substitution with individuals that could infect others (a risk reduced by combining serology with PCR).

Thus far we have focused on the impacts of shield immunity as a singular strategy, yet in practice, multiple interventions will be used in parallel. Hence, we evaluated the synergistic potential of utilizing shield immunity in combination with social distancing. Social distancing is modeled as a reduction in the transmission rates sustained over the post-intervention period. As is apparent in Figure 4, shielding can augment social distancing, particularly when social distancing is relatively ineffective. For example, contour lines of reduction in total fatalities suggest that a combination of 10% reduction in transmission with *α* = 20 is equivalent to a nearly 50% reduction in transmission in the absence of shield immunity. However, there is a trade-off. Because social distancing reduces contacts and transmission, there are fewer recovered individuals when *β* is reduced by 50%. Nonetheless benefits of shielding accrue at all levels of social distancing. Social distancing and shield immunity may work in combination to improve outcomes in terms of expected hospitalization burden, again suggesting a role for shield immunity in reducing transmission and reducing the negative impacts of suppression-level social distancing policies. Finally, we note that targeted shield immunity may also enhance population outcomes by focusing the effort of recovered individuals in subsets of the population. In the SI, we show heuristic solutions to an optimization formulation of targeted (i.e., age-specific) shield immunity in this model. In effect, by preferentially targeting older individuals, it is possible to further reduce cumulative deaths by ≈30% (see Figures S9-S11).

**FIG. 4:**
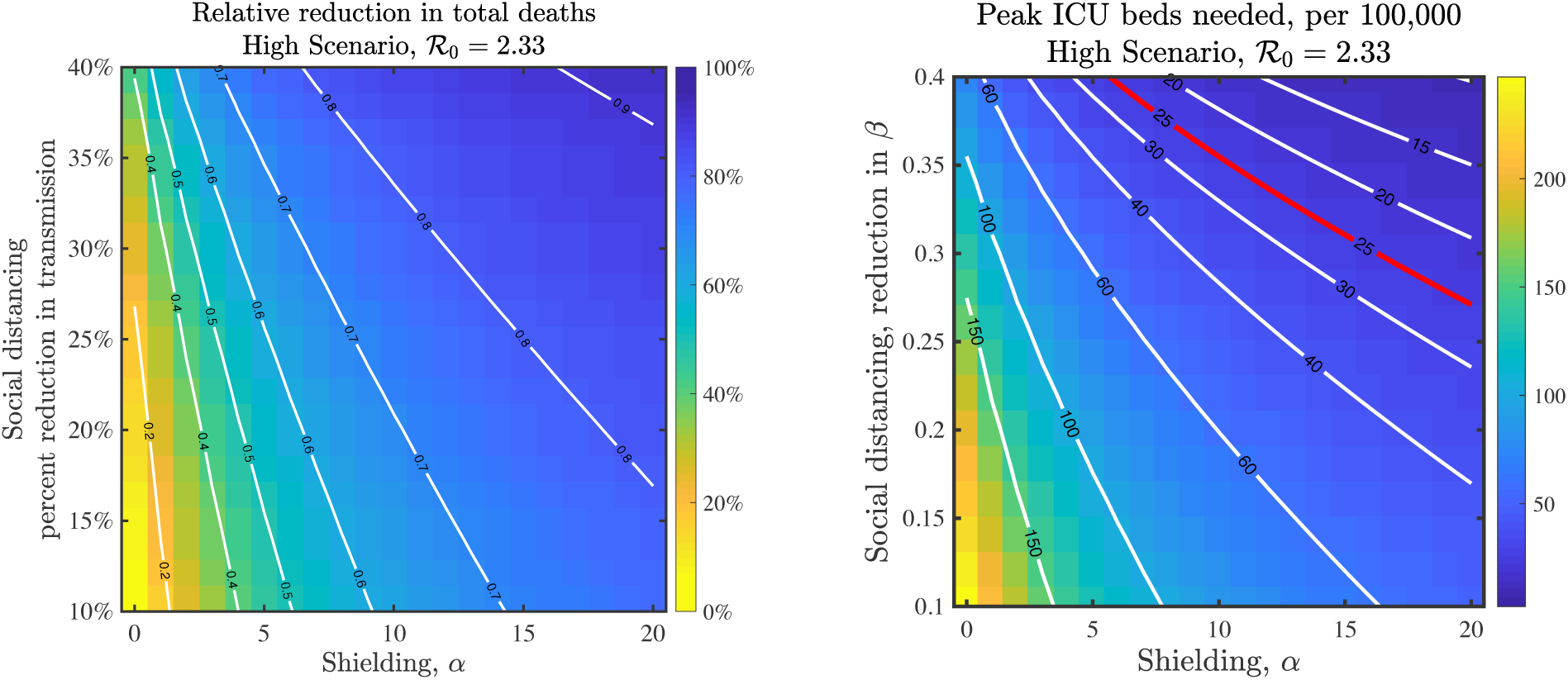
Impacts of combined interventions of shielding and social distancing in the high scenario. (Left) Fractional reduction in deaths compared to baseline; (Right) Peak level of ICU beds needed on a given day during the epidemic; the red line denotes 25 ICU beds per 100,000 individuals as a demarcation point for surge capacity.

Serology testing is needed now, at scale, for many reasons. Here, we have shown a rationale for serology testing as a means to facilitate interventions beyond those of mitigation and suppression. Identifying and deploying recovered individuals could represent more than just a metric of the state of the COVID-19 epidemic, e.g., to better measure prevalence and the ‘denominator’, but an opportunity to slow transmission by developing population-level shield immunity. Many logistical, social, and dynamical challenges remain if such an idea were to move from theory to feasibility. Accurate and rapid serological tests are needed at scale, including targeted surveys to identify essential workers and via population-level surveys. The potential scale of shield immunity depends on both the intrinsic epidemic dynamics, driving the number of recovered individuals able to provide shield immunity, and also on the ability to identify and deploy them (e.g., via the shielding parameter *α*). Yet, even if such tests were available, who should get them? Public health authorities and governmental agencies should consider how to prioritize those in critical roles, those with experience in disaster response, as well as prior individuals who have tested positive for COVID-19 (and could then return for both serology-based and viral shedding assays). Positive confirmation of immunity and cessation of viral shedding could help identify and deploy (tens of) thousands of individuals as part of a shield immunity strategy, with the greatest concentration likely co-located with areas in greatest need of intervention. A national (or global) strategy could consider the deployment of critical response workers to help control new outbreaks.

We recognize there are significant challenges to developing and implementing interventions that aim to develop population-wide shield immunity. Nonetheless, the magnitude of the current public health and economic crisis demands large-scale action (e.g., [33, 34]). The efficacy of a shield immunity strategy will be demographic, test-and context-dependent. Beyond the near-term, the duration of immune memory is also relevant in projecting to a multi-year post-pandemic framework where demographic dynamics and strain evolution are increasingly relevant [35, 36]. In moving forward, it will be critical to understand how shield immunity is modulated by spatial and network structure. In a network, well-connected individuals have a disproportionate effect on the spread of disease [20]. Network structure represents an opportunity to position immune shields at focal points of ‘essential’ services, and even to prioritize the focus of population-scale serological prevalence assays based on the connectivity of targeted individuals. Although the number of laboratory confirmations is both staggering and growing, the actual number of infections is higher – likely far higher; for example in China, 80% of transmission of new cases were from undocumented infections [26] and there is significant uncertainty with respect to case ascertainment [37]. Asymptomatic transmission may paradoxically provide a greater pool of recovered individuals to develop shield immunity at scale. We contend that it is time for collective action to ascertain more information on prevalence and to consider strategic use of serology as the basis for public health intervention to slow the pandemic spread of COVID-19.

## Data Availability

All simulation and codes used in the development of this manuscript are available at https://github.com/WeitzGroup/covid_shield_immunity.

## Acknowledgements

We thank P.S. Dodds, A. Kraay, M. Lipsitch, K. Levy, B. Lopman, D. Muratore, K. Nelson, A. Sanz, and J. Shaman for comments and feedback. Tweet threads by T. Bedford and N. Christakis were influential in refining initial ideas into the current form. Research effort by JSW and co-authors at the Georgia Institute of Technology was enabled by support from grants from the Simons Foundation (SCOPE Award ID 329108), the Army Research Office (W911NF1910384), National Institutes of Health (1R01AI46592-01), and National Science Foundation (1806606 and 1829636). JD was supported in part by grants from the Canadian Institutes of Health Research and the Natural Sciences and Engineering Research Council of Canada.

